# The effect of BMI and physical activity levels on the duration of symptomatic days with Covid-19 infection

**DOI:** 10.1101/2020.08.21.20179499

**Authors:** Abdulazeem S Alataibi, Boukhemis Boukelia

## Abstract

Regular exercise is known to boost immunity, increase immune response to fight infection, as well as speeding up recovery times and healing processes. This study seeks to assess if exercising regularly pre-SARS-CoV-2 (COVID-19) and/or BMI status has an effect on recovery time.

A total of 215 people infected with COVD-19 from the Kingdom of Saudi Arabia took part in this study (age 36±16 years, mass 72±15 kg, stature 166±11 cm). Only 10 patients were physically active and fulfil WHO physical activity requirement (Age 30±7 years, Mass 77±9 kg, Stature 176±1 cm).

There was a significant difference in recovery time between active and inactive patients (P = 0.00) with active patients’ recovery 2.7 times faster than inactive patients. Active patients showed a lower BMI level (p = 0.043).

Anthropometric measurement characteristics and the fitness level could be used in decision making scenarios for the estimation of the risk of complications in patients with COVID-19.

**Novelty:**

- Covid-19 physical active patients shows faster recovery time.
- Active patients recorded a BMI of over 25kg/m^2^, recovered faster than those inactive patients with similar BMI’s.

## Introduction

The rapid spread of the corona virus that causes COVID-19 (SARS-CoV-2) has sparked alarm worldwide. The way the virus spreads is still debated; however, it is known that similar viruses are spread in cough and sneeze droplets and possibly in aerosols. At the time of writing this manuscript over eleven million people worldwide have been infected with over five hundred thousand deaths (WHO, 2020). Symptoms of COVID-19 are often mild and begin gradually, and include fever, fatigue, dry cough, some pain and aches, nasal congestion, cold, sore throat, and diarrhea (Zheng et al., 2020), whereas some people become infected without showing any symptoms at all. The prevalence of developing COVID-19 symptoms among populations with chronic disease are high (Pablos et al., 2020). In Garg et al. (2020), among patients aged 18–49 years, obesity was the most prevalent underlying condition, followed by asthma and diabetes mellitus. In patients over 50 years, obesity was most prevalent, followed by hypertension. It has also been reported that COVID-19 is more likely to occur in older and adults’ people within weaker immune functions Chen et al., 2020).

It is well documented that physical activity stimulates the human immune system and strengthens the infection defence system (Simpson et al., 2006). Moreover, evidence suggests that sedentary people who become physically active get a progressively stronger immune system and become less susceptible to infections (Nash, 1994). Hence, many studies support the concept that engaging in regular bouts of moderate aerobic exercise protect against respiratory infections (Nieman et al., 1993; Nieman et al., 2011; Boukelia et al., 2016). For example, the risk of upper respiratory tract infection reduced by up to 67% compared to sedentary population (Gleeson, 2007) Outstandingly, 20–40 minutes of moderate daily exercise was reported to be adequate to promote a beneficial effect on the immune system. This exercise-induced protection against infection can be achieved with many types of aerobic activity such as running, swimming, walking, and dancing (Nieman et al., 2011). The mechanism behind the moderate exercise protection against respiratory infection remain debated. Nonetheless, there are several possible reasons to explain this, such as, an increase in natural killer cells, neutrophils, and antibodies in the blood stream (Miles et al., 2002). For each bout of exercise, the human body mobilizes billions of immune cells types that are capable of recognising and killing virus-infected cells. In addition, exercise also releases muscle-derived cytokines which help to maintain immunity such as IL-6, IL-7, and IL-15. IL-6 has been shown to mobilise immune cell to the infected area, while IL-7 helps to maintain the peripheral T-cell and NK-cell compartments which assist to increase the human body resistance to infection (Simpson, 2020). Furthermore, improved psychological well-being could be another possible factor (emotional stress control).

The Kingdom of Saudi Arabia has been reporting an average of 11.1 cases per 100,000 people with a mortality rate of 0.91% (MOH 2020). WHO ranks the outbreak of COVID-19 at the orange risk level (the second highest risk category) which means it is quickly spreading (WHO, 2020). A preventative measure for reducing the likelihood of this or future infections may be found through increased activity and reduced obesity within the population.

Three quarters of the adult population are classified as inactive in KSA (Al-Hazzaa, et al., 2011), following similar trends of developed countries across the world. Nevertheless, this sedentary lifestyle has been associated with a greater risk of early mortality from obesity related diseases that equate to 6% of the total death rate globally each year (Henriksen et al., 2019). This study aims to investigate the effect of fitness level on the rate of recovery time among positive COVID-19 tested in KSA.

## Methodology

A questionnaire developed by the Health and Exercise Science Department at al Qassim University, KSA were used in this study. A 12 multiple choice questionnaire using a self-rated scale was used to evaluate the effect of fitness level on the recovery time in SARS-CoV-2 (COVID-19) patients. 215 (mean age ± 36.3±16.2) COVID-19 infected participants from the region of Riyadh, KSA took part in this study. All infected patients observed self-isolation within medically monitored hotels and kept away from their families to minimize the risk of spreading the virus. All patients were under direct supervision of a medical team and received daily assessment on their state of health, patients that tested negative for the virus were allowed to return home. The participants were divided into two groups, a sedentary group and an active group that fulfills WHO activity criteria (no less than 150 minutes of physical activity a week). Analysis of the data revealed that from a total of 215 patients only 10 patients exercise routinely pre-positive test and fulfill the activity recommendation by WHO (150 mins a week). Body mass, age, ethnicity, diet, and sleep level were recorded as well.

All participants provided fully informed written consent before engaging with the experiment. This study was approved by the Al Qassim University Ethics Committee and was conducted in accordance with the guidelines of the 1964 Declaration of Helsinki and its later amendments or comparable ethical standards.

### Statistical Analysis

Prior to statistical analysis, all data were checked for normality. Repeated measure T tests were used to determine significance difference in Active and Inactive patients and BMI in both groups (SPSS20 Statistical Software, IBM.UK). Statistical significance was accepted at P < 0.05. Results are represented as mean values ± standard deviation (SD). Cohen’s d was used to assess the size of the difference between two related sample.

### Results

**Table 1:** Anthropometric measurements

| Measurements | Physically Active | Physically inactive |
| --- | --- | --- |
| Age (years) | 30±7 | 34±6 |
| Stature (cm) | 176±0.1 | 170±0.06t |
| Mass (kg) | 77.8±9.75 | 78.8±7.6 |

**Table 2:** The average sleeping hours pre and during the quarantine period.

| Sleeping hrs | Active (%) |  |  | Inactive (%) |  |  |
| --- | --- | --- | --- | --- | --- | --- |
|  | Pre-quarantine | During quarantine | Δ | Pre-quarantine | During quarantine | Δ |
| >5 hrs | 9.7 | 19.4 | -9.7 | 14 | 15.4 | -1.4 |
| 5-7 hrs | 19.4 | 18.1 | +1.3 | 16.8 | 21 | -4.2 |
| <7 hrs | 70.8 | 62.5 | +8.3 | 69.2 | 63.6 | +5.6 |

There was a significant BMI difference between active and inactive patients t (19) = 2.15, p = 0.043 (graph 1).

**Graph 1:**
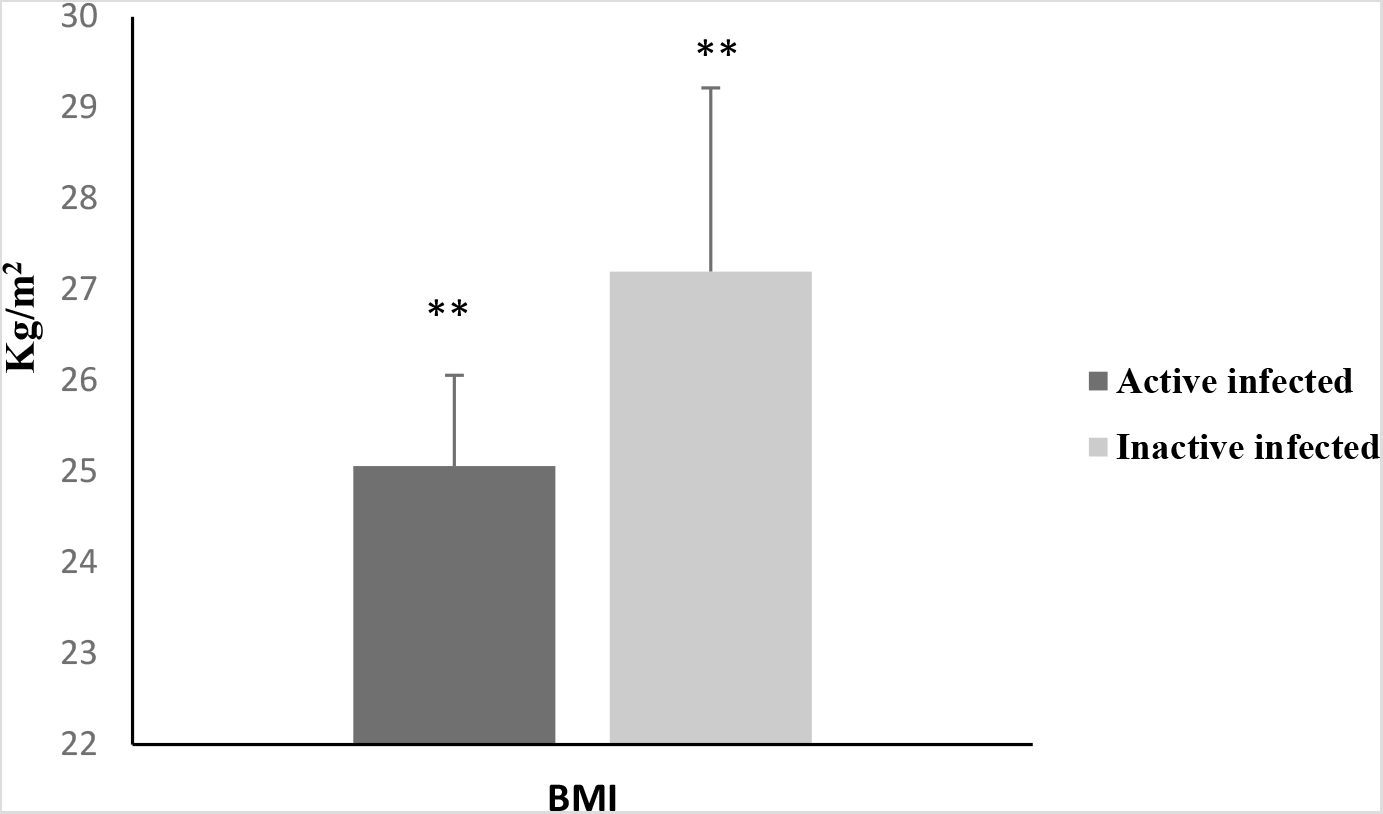
Active vs inactive tested Covid-19 positive BMI. ** denotes a significant different between active and inactive patients BMI (P < 0.05), values are mean ± SD.

Inactive patients shows a significant slower recovery time compared to active patients t(19) = 3.29, p = 0.004 (graph 2). Active patients were recovered 2.7 times faster from COVID-19 infection than the group of inactive infected patients.

**Graph 2:**
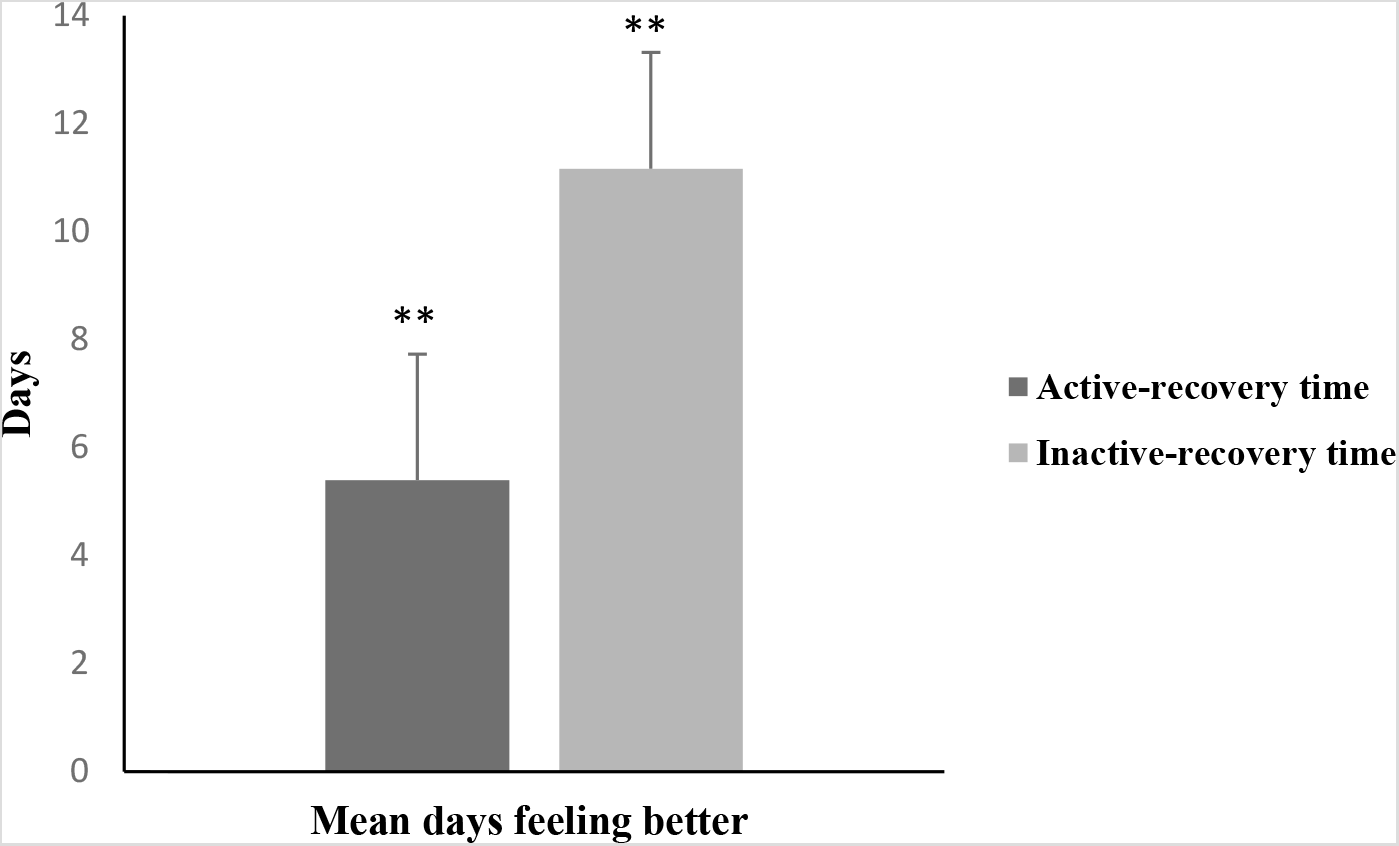
the mean days of feeling better active vs inactive infected patients. **denotes a significant different between active and inactive patients in the time of feeling better (P < 0.05), values are mean ± SD

### Discussion

To date this is the first study to assess the effect that the level of physical activity and recovery period on COVID-19 infected patients. Therefore, this study provides evidence-based insights on the relationship between inactivity, higher BMI, and COVID-19 recovery time. Only 4% of the patients in this study were physically active in either amateur sport or recreational physical exercise that fulfil the WHO recommended level of activity requirement. A significant difference in BMI between active and inactive patients were evident in this study and has been shown to have a significant difference on the duration of recovery. the recovery period for the inactive population was slower by 207% compared to active patients. The outcome of this study supports the finding that individuals with higher BMI are more likely to have respiratory symptoms than individuals with a normal BMI (Gibson, 2000; El-Gamal et al., 2005; Babb et al., 2008). In a new report revealed by PHE, (2020) patients with over 25kg/m^2^ of BMI (clinically overweight) are more than three times more likely to die of COVID-19 and seven times more likely to need a ventilator. In Stefan et al. (2020) 85% of patients with higher BMI required mechanical ventilation and 62% of the patients with higher BMI died of COVID-19, compared with 36% of those with a healthy weight.

Despite meeting the WHO recommended activity levels 4 out of 8 active patients in this study recorded a BMI of over 25kg/m^2^ BMI of over 25kg/m2, nevertheless, they likewise recovered faster than those inactive patients with similar BMI’s (an average of 3.75 days vs 10 days). Therefore, it has reported that increasing physical activity including those currently overweight, can reduce the risk of cardiovascular disease and improve health by being physically active (Koolhaas et al., 2017). The average participant age from this study of 215 was 36±16 years old. Therefore, this study would support the findings of Kass et al. (2020) that found a significant inverse correlation between age and BMI, in which younger individuals that are overweight are least severely affected by COVID-19 (Kass et al., 2020). Saudi Arabia has a rising obesity problem with approximately 40% of the total population categorised as obese Alqarni, 2016). Therefore, this phenomenon may contribute to the fast spread and increased symptom severity and/or duration of COVID-19; that would consequently greatly stress the national health service.

It must be considered that the low proportion of physically active patients recorded in this study may in fact be linked to asymptomatic infections that have remained undetected in the general population. Therefore, measurement of anthropometric characteristics and the level of activity could be key to estimating the risk of complications in patients with COVID-19. In addition, countries with high overweight or obese populations should adequately characterise the risk obesity poses in relation to COVID-19 symptoms alongside other contributing factors or chronic illnesses; particularly when considering future vaccination programs on the basis of risk. Individuals carrying excess weight will have poor responsiveness to vaccination against COVID-19 due to the impaired activation and function of t cells adaptive immune cells (Green and Beck, 2017), however, the development of novel immunization strategies for this population is warranted.

## Data Availability

The redacted datasets generated and analysed during the current study are available from the corresponding author on request

## Tables

The mean age, stature and mass were presented in table 1. Active patients showed 4 years younger than inactive patients. In addition, this data shows a correlation between age and physical activity.

Active patients showed better sleeping hours compared to inactive patients at pre the quarantine period (Table 2). Whereas, during the quarantine period 19.4% of active patients sleep less than 5 hours compared to 9.7% at pre-quarantine period.

There was a significant difference between active and inactive patients BMI t (19) = 2.15, p = 0.043 (graph 1).

